# Cohort Profile: Immune Responses to SARS-COV-2 Vaccination and Infection in a Longitudinal Sampling Amidst the COVID-19 Pandemic (LONGTONG-SARS2) in Malaysia

**DOI:** 10.1101/2024.03.26.24304850

**Authors:** Naim Che-Kamaruddin, Jefree Johari, Hasmawati Yahaya, Huy C. Nguyen, Andrew G. Letizia, Robert D. Hontz, Sazaly AbuBakar

**Author notes:** **Email addresses:** Naim Che-Kamaruddin, Jefree Johari, Hamawati Yahaya, Huy C. Nguyen, Andrew G. Letizia, Robert D. Hontz, Sazaly AbuBakar.

## Abstract

**Purpose:** This prospective, longitudinal study aims to evaluate the durability and functionality of SARS-CoV-2 Ancestral strain (Wuhan-Hu-1)-specific immune responses induced by COVID-19 vaccination and natural infection over a 12-month period. This article reviews the study protocol, design, methodology, ongoing data collection, analysis procedures, and demographic characteristics of the cohort enrolled.

**Participants:** Between March 2021 and May 2022, 400 participants were enrolled with a 12-month follow-up, concluding in May 2023. Two main groups of participants: (1) serologically SARS-CoV-2-naïve individuals receiving the BNT162b2 primary series vaccination (referred to as VAC) and (2) those who recently recovered from COVID-19 infection within 30 days, regardless of vaccination history (referred to as COV). Additionally, a subset of 45 participants with selected COVID-19 exposure histories provided peripheral blood mononuclear cells (PBMCs) for cross-sectional analysis six months after enrollment.

**Findings to date:** Out of 400 participants, 66.8% (n=267) completed the follow-up. Among them, 52.8% (n=141) were in VAC, and 47.2% (n=126) were in COV. As the study progressed, we acknowledged cross-over between initial groups, leading to restructuring into five revised groups based on sequential exposure events. Sociodemographic factors revealed statistically significant age distribution differences (p=0.001) in both initial and revised groups, with no significant differences observed for sex.

**Future plans:** LONGTONG-SARS2 assesses the host-pathogen interactions central to the development of COVID-19 immunity. With enrollment spanning two years of the pandemic, most participants exhibited mixed SARS-CoV-2 exposures—via vaccination and infection—resulting in diverse subgroups of interest. Notably, the inclusion of SARS-CoV-2-naïve, pre-exposure serum samples allowed for robust comparator and reduced potential biases. Ongoing analyses will include serology kinetics, memory cells ELISpots, B cells repertoire analysis, cytokine/chemokine profiling, and proteomic pathway to comprehensively examine the immune response against the SARS-CoV-2, thus informing and potentially predicting dynamic longitudinal responses against new more transmissible, immune-evasive SARS-CoV-2 variants.

**STRENGTH AND LIMITATIONS OF THIS STUDY:**

- LONGTONG-SARS2 is a prospective longitudinal study that comprehensively evaluates the SARS-CoV-2 immune response among a diverse group of individuals, stratified based on the sequential order of SARS-CoV-2 exposure events, whether from COVID-19 vaccination or infection.
- Pre-vaccination serum samples were collected from serologically SARS-CoV-2 naive individuals scheduled to receive the BNT162b2 primary series vaccination during the initial mass COVID-19 vaccination phase in Malaysia in early 2021.
- The longitudinal serum sample collection spanned two years of the COVID-19 pandemic, from March 2021 to May 2023. This extended duration allows for robust monitoring of the immune response against SARS-CoV-2 variants in comparison to the ancestral strain.
- There is a risk of misclassification of some individuals’ SARS-CoV-2 exposure status through serology, as certain sampling timepoints had intervals of three months. Additionally, our study relies on self-reported data through the *MySejahtera* application (Malaysia’s electronic medical record by the Ministry of Health) for second confirmation, potentially leading to underdiagnosed and underreported cases of asymptomatic infection.

## 1.0 INTRODUCTION

Coronavirus disease 2019 (COVID-19) is an ongoing viral respiratory illness caused by Severe Acute Respiratory Syndrome Coronavirus 2 (SARS-CoV-2). SARS-CoV-2 is a positive-sense, single-stranded RNA virus. This Risk Group 3 (RG3) virus is phylogenetically related to other Betacoronaviruses which can cause respiratory illnesses in humans, including Severe Acute Respiratory Syndrome Coronavirus 1 (SARS-CoV-1) and the Middle East Respiratory Syndrome Coronavirus (MERS-CoV) [1]. As of 31 December 2023, SARS-CoV-2 has been responsible for more than 773 million COVID-19 cases and more than 7 million deaths globally, with the actual death toll likely to be higher, largely due to the lack of reliable mortality surveillance and tracking systems in many countries and instances where individuals succumbed to COVID-19 before being tested for the virus [2]. Most patients infected with SARS-CoV-2 experienced asymptomatic, mild, or moderate symptoms, and recovered without hospitalization, while a minority developed severe or critical clinical manifestations [3].

The effectiveness of SARS-CoV-2 vaccines has been well documented [4–6]. However, waning antibody titers which has been documented following three to four months post-infection and reports of reduced vaccine efficacy against emerging SARS-CoV-2 variants underscore the need for ongoing assessment of continuous booster immunization [7]. Previous studies suggested that the risk of reinfection is attributed to the waning of neutralizing antibodies (NAb) [4–7]. According to these earlier studies, antibodies are only detected in 60–85% of convalescent individuals, despite detectable levels in 80–95% within two weeks post-symptom onset [8–10]. Considering the serologic kinetics, booster administration increased the serum NAb titer and cross-reactivity against emerging Omicron variants [11]. Although booster immunization has been shown to stimulate antigen-specific antibody production, additional more comprehensive studies are needed to better understand the multifaceted immunological mechanisms that correlate with clinical protection following vaccination and/or natural infection, particularly in the context of the continuing emergence of new immune-invasive SARS-CoV-2 variants [12].

SARS-CoV-2 exposure from vaccination and/or infection elicits an adaptive immune response, engaging both humoral (primarily antibody-mediated) and cellular immunity, which includes the activation of B and T cells [13]. This dual response establishes long-term memory within the immune system. Upon re-exposure to SARS-CoV-2, the memory B and T cells expand to mount a rapid and targeted defense [6]. Immune memory cells, demonstrated to be more durable, contribute to long-term immunity against variants, even as antibody titers contract [5,6,14]. Evidence of immune imprinting, wherein initial exposures effectively prime B cells but potentially limit the development of novel memory B cells and NAb against emerging SARS-CoV-2 variants, poses a significant challenge to maintaining vaccine efficacy amid the ongoing emergence of novel SARS-CoV-2 variants [15,16]. While the mechanisms of how the immune system recognizes and responds to SARS-CoV-2 variants are still being elucidated, both humoral and cellular immunities play important roles in conferring protection [17]. Therefore, it is imperative to investigate the impact of varied SARS-CoV-2 exposure histories on shaping the immune responses to SARS-CoV-2 [16], especially on a regional basis. This is attributed to the variation in immune response across population, with factors such as genetic, environmental, and lifestyle being unique to each region [18]. Moreover, the prevalence of SARS-CoV-2 variants and vaccination strategies in each region can significantly impact viral transmission dynamics. This information becomes crucial for informing public health policies, particularly in the context of emerging highly transmissible variants [19].

The present study seeks to establish a comprehensive longitudinal prospective cohort comprised of participants exposed to SARS-CoV-2 vaccination and/or infection to define the epidemiological and immunological effects of SARS-CoV-2 in Malaysia. To our knowledge, Malaysia lacks a cohort with a similar extensive design, involving longitudinal sampling up to 12 months, spanning from the early phase of the pandemic to the transitioning to an endemic phase. This study fills a crucial gap by providing insights into COVID-19 humoral and cellular immune responses specific to Malaysia. Through investigation of antibody kinetics, T and B cell studies, and adaptive immune responses, our initiative not only contributes valuable data for informing public health strategies but also has the potential to serve as a reference for future research in the region. This article describes the protocol and cohort profile of **LONG**itudinal s**T**udy **O**f immu**N**e response a**G**ainst SARS-CoV-2 (LONGTONG-SARS2).

### 2.0 STUDY OBJECTIVES

The overall aim of “**LONG**itudinal s**T**udy **O**f immu**N**e response a**G**ainst SARS-CoV-2” (LONGTONG-SARS2) is to evaluate the durability of SARS-CoV-2 specific antibodies, and the functionality of the specific memory B and T cells from a prospective, longitudinal sampling for up to 12 months. The specific objectives include:

1. To analyze the kinetics of anti-SARS-CoV-2 antibodies (IgM, IgA, IgG, NAb) response for up to 12 months by ELISA assays for each group.
2. To assess the functional immune memory response of SARS-CoV-2-specific memory B and T cells by ELISpot assays.
3. To characterize the interaction of cytokines and immune cells through bead-based analytes.
4. To profile the serum protein pathways before and after SARS-CoV-2 vaccination or infection from bottom-up proteomic analysis.
5. To identify the distribution and incidence of SARS-CoV-2 exposures by duration since the primary series of vaccination doses, frequency of infection(s), and date of exposure in the study population by descriptive analysis.
6. To evaluate the association of antibody distribution and baseline demographic, comorbidities, as well as symptoms/adverse effects following immunization (AEFI) observed during enrollment, subsequent to SARS-CoV-2 exposures via vaccination or infection by generalized estimating equation (GEE) analysis.

### 3.0 COHORT DESCRIPTION

### 3.1 Terminology definitions

The terminology used in the present study are:

- "Vaccination" - Participants were classified as vaccinated if they received COVID-19 vaccination regardless of number of doses. Proof of vaccination is a mandatory requirement for inclusion in any vaccination data collection within the scope of the present study. Participants who received the primary vaccination series were categorized as fully vaccinated two weeks after the last dose, while those receiving additional vaccine doses after the primary series were classified as boosted.
- "Infection" - Participants were classified as infected if they presented proof of a positive case through PCR or rapid test results in either their records (as well as from the electronic medical record by Ministry of Health, *MySejahtera* application) OR demonstrated at least a 4-fold increase in IgG or IgM levels from the consecutive sampling timepoints. Individuals exhibiting a 4-fold increase in IgG or IgM without documented proof (due to the absence of symptoms) were considered to have asymptomatic infection.
- "Reinfection" - Reinfection status for participants was documented if multiple occurrences of COVID-19 infections were recorded in the electronic medical record OR if there was at least a four-fold increase in IgG or IgM levels from the consecutive sampling timepoints even without the records.
- "Breakthrough infection (BTI)" - The BTI status for participants was documented if the date of infection occurred after Day-15 of the second dose of COVID-19 vaccination, indicating completion of the vaccination regimen.
- “Exposure” –Vaccination, infection, reinfection, and BTI were considered as exposure in the present study.
- "Cross-exposure" - Participants were defined to have cross-exposure if they experienced both COVID-19 vaccination and infection, irrespective of the sequence of exposure.

### 3.2 Study design

The study design involved a longitudinal study with enrollment of two groups of different SARS-CoV-2 exposures, which are (1) the serologically SARS-CoV-2-naïve individuals who were scheduled to receive primary series vaccination with BNT162b2 (VAC) and (2) post-COVID-19 infected group (COV) regardless of their vaccination histories. The participants were informed beforehand about all aspects of the study, and individuals who fulfilled the inclusion criteria were given an informed consent form and a case report form (CRF) adhering ethical approval (MREC-UMMC SID: 2021226-9886).

The inclusion criteria are as follows:

- Seven years or older, AND
- Healthy individuals without active SARS-CoV-2 infection at the time of enrollment, AND
- Healthy individuals without immunosuppressive conditions such as HIV and cancer patients, AND EITHER
- Scheduled for the primary series vaccination with BNT162b2 with a negative baseline SARS-CoV-2 anti-S1 IgG serology, corresponding to no previous SARS-CoV-2 exposure (to be enrolled in the VAC group), OR
- Had been infected with SARS-CoV-2 within 30 days and able to provide proof of infection either from rapid test kit or PCR (to be enrolled in the COV group),
- AND agreed to give voluntary informed consent to participate in the study

The exclusion criteria are as follows:

- Individuals with immunosuppressive conditions such as HIV and cancer patients, OR
- Individuals unable to attend the scheduled follow-up appointments

Epidemiological data obtained in the CRF include:

- Demographics - *Age, gender, body mass index (BMI), medical comorbidities*
- Exposure history - *Date of vaccinations, COVID-19 vaccine types, date of COVID-19 infections*
- Clinical manifestations experienced - *Adverse effect following immunization (AEFI) and symptoms following COVID-19 infection*

### 3.3 Sample size estimation

A sample size of 400 participants (N=400) was targeted to achieve statistical power as calculated using GLIMMPSE software (https://glimmpse.samplesizeshop.org/), taking into consideration the repeated measures and longitudinal design [9,10,20].

In GLIMMPSE, the Hotelling-Lawley Trace test was utilized with a conservative Type 1 error rate of 0.5, with the total sample size for the analysis being 66 participants. An estimated 30% attrition rate was used, and an adjusted estimation formula of 66 / (1-0.3) was applied to accommodate this attrition, resulting in a minimum total sample size requirement of 95 participants. For the present study, the total of 400 participants were chosen to increase the statistical power and accommodate potential loss of follow-up participation.

### 3.4 Enrollment

Following all required ethics approvals, eligible participants were enrolled. Enrollment material (i.e., advertisements, posters, and flyers) were posted on social and printed media. The target study participants primarily comprised of individuals from the public residing in the greater Kuala Lumpur area (Supplemental Figure 1). Due to the restrictions imposed by the nationwide MCO, we utilized both convenience sampling, selecting participants based on their accessibility and proximity to the study site, and snowball sampling, where existing participants referred others for the study. Participant enrollment commenced in March 2021, coinciding with the initial phase of COVID-19 vaccine distribution in Malaysia. Initially, potential participants were provided with comprehensive information on the study, which included the study purpose, blood collection procedures, and the prerequisite follow-ups. Consented participants completed informed consent. Visit schedules for sample collection were sent via an electronic appointment card. Collected blood samples were transported on ice to the laboratory, where the serum samples were processed and stored in -80°C freezer until needed for laboratory analysis.

Enrollment closed at the end of July 2022, and the blood sampling ended at the end of May 2023.

### 3.5 Blood sample collection

Longitudinal blood sampling for up to 12 months was required for all participants following the timepoints described in Figure 1. Participants in the VAC group were scheduled for a total of 9 sampling timepoints, spanning from pre-vaccination or baseline to Day-360 days post the first vaccine dose. Meanwhile, the COV group had 7 scheduled timepoints, initiated from the first month of infection. Notably, sample collection timepoints for both groups coincided on Day-14 post first exposure, Day-90, Day-180, Day-270, and Day-360 post-vaccination/infection. A sampling window of ±7 days was applied to the designated sampling timepoints for participants’ flexibility and availability. One (5 mL) tube with a separator gel and clot activator for whole blood samples was drawn for each timepoint. Blood samples were processed with serum extracted from each sample for the serology testing.

**Figure 1.**
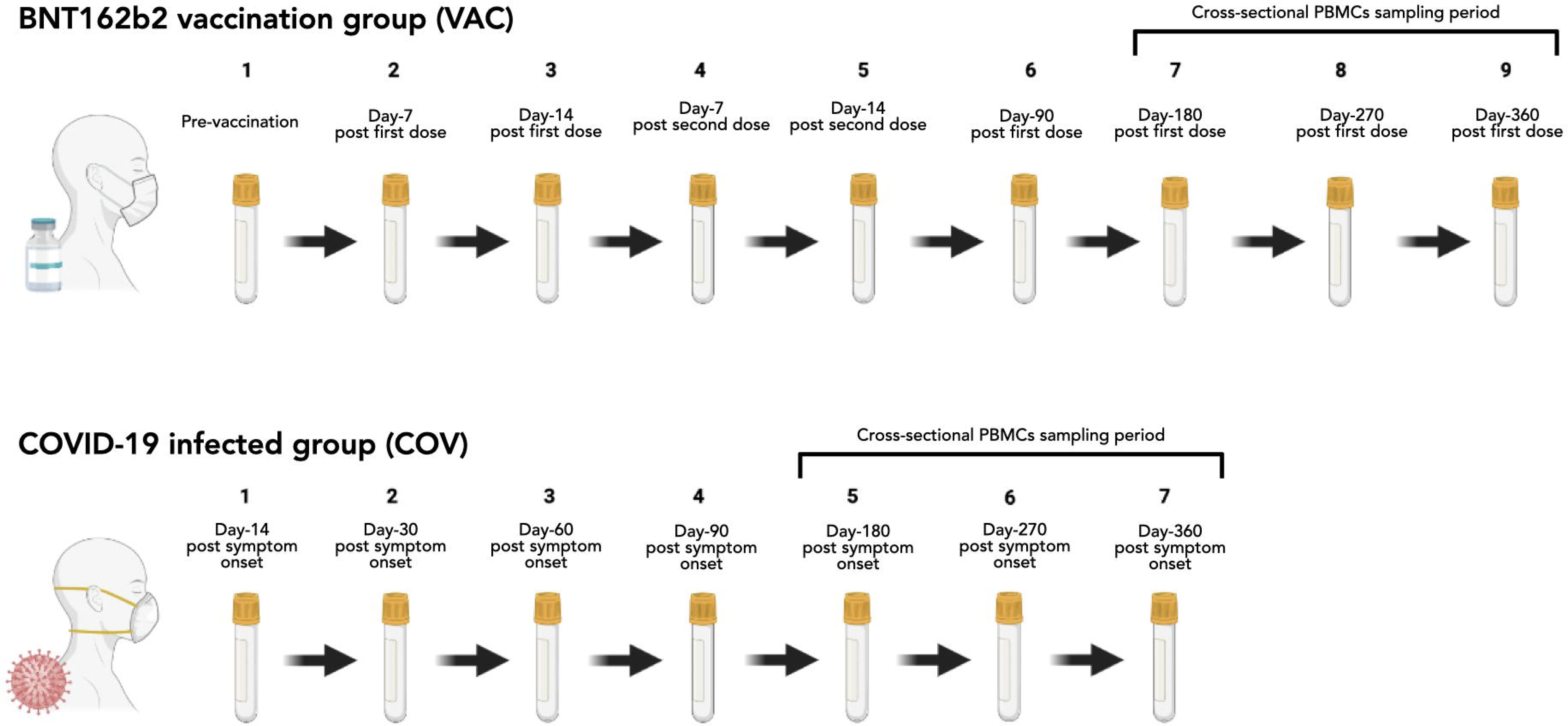
Schedule of longitudinal blood sampling collection for up to 12 months and the cross-sectional PBMCs sampling period during the expected waning antibody phases.

A cross-sectional collection of Peripheral Blood Mononuclear Cells (PBMCs) was conducted from Day-180 to Day-360 to assess the cellular immune response. Among the 400 participants, 45 were randomly selected and consented for PBMCs collection. To establish a control group for cellular immune response analysis, an additional 5 participants who had no COVID-19 vaccination documented in a local electronic record, *MySejahtera* and were not previously involved in the LONGTONG-SARS2 serology study, were included. Details of the vaccination types and the frequency of SARS-CoV-2 infection were collected to form groups based on these parameters (details are shown in Supplemental Table 3). For the PBMCs collection, four (8 mL) lymphocyte isolation tubes (BD Vacutainer® CPT™ Mononuclear Cell Preparation Tube) were used. The PBMCs were isolated and assessed for cell count estimation and viability using BioRad TC20™ Cell Counter.

### 3.6 Participant and public involvement

Participants or the public were not involved in the design, or conduct, or reporting, or dissemination plans of our study.

### 4.0 LABORATORY INVESTIGATIONS

There are five major components of laboratory analysis: i) serology monitoring, ii) B- and T-memory cell functionality, iii) B lymphocyte population counting, iv) cytokine profiling, and v) proteomic profiling (Figure 2). Briefly, the serology monitoring involves the longitudinal quantification for specific antibodies against SARS-CoV-2. The assessment of B- and T-memory cells functionality seeks to understand how well the immune memory responds upon re-exposure to SARS-CoV-2 antigens. B lymphocyte population counting intends to elucidate the heterogeneity in B cells population following the re-exposure, while cytokine profiling involves identifying key signaling molecules that are upregulated or downregulated as part of the immune response. Additionally, proteomic profiling provides a broader view of protein expression patterns during the immune response. Profiling the various arms of the immune system will help establish a comprehensive analysis for a holistic understanding of how the immune system responds against SARS-CoV-2.

**Figure 2.**
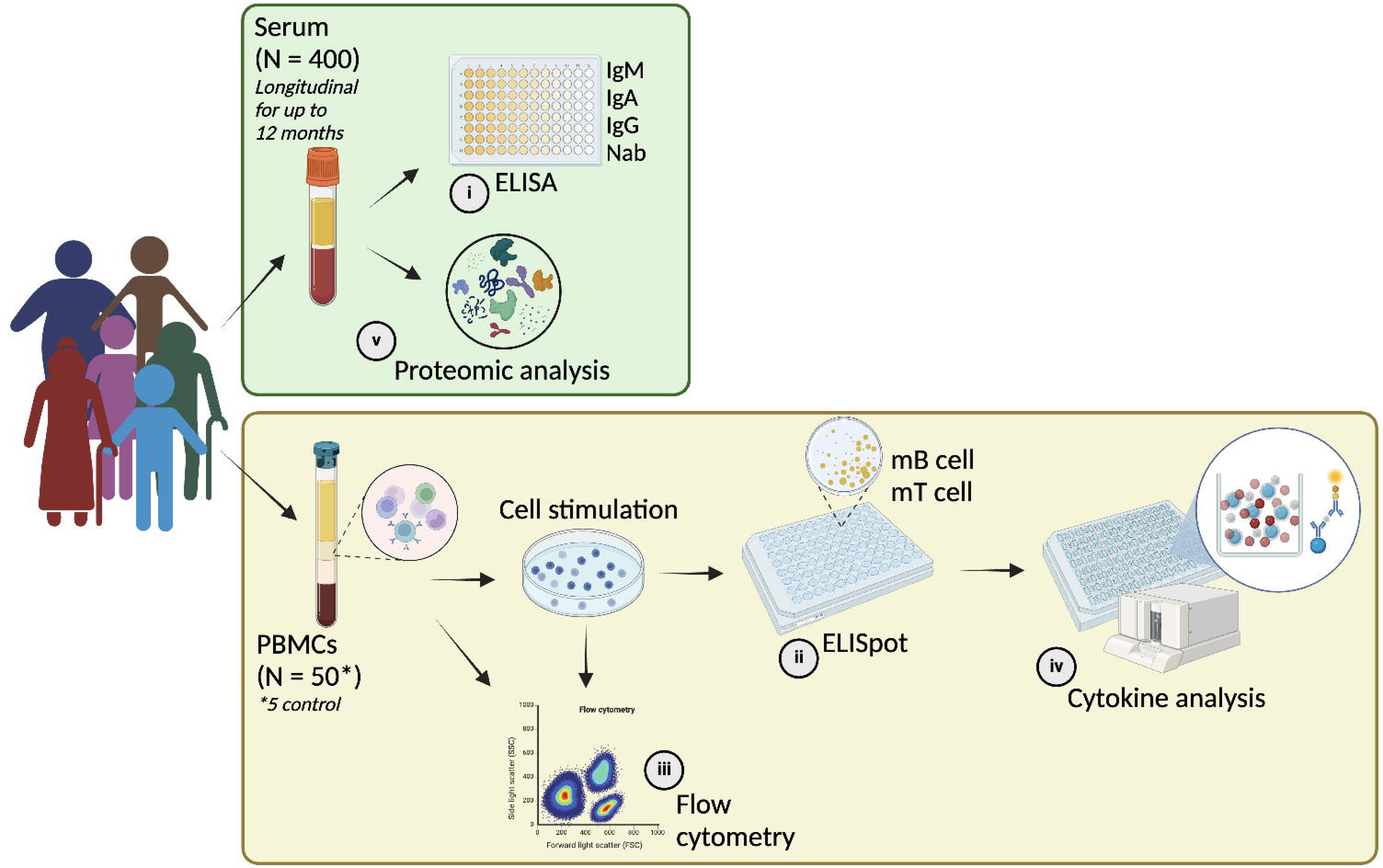
Overall LONGTONG-SARS2 study design of laboratory analysis comprising of i) serology monitoring, ii) B- and T- memory cell functionality, iii) B lymphocyte population counting, iv) cytokine profiling, and v) proteomic profiling.

### 4.1 Serology monitoring

Serological testing of serum samples employs four different commercially available anti-SARS-CoV-2 ELISA kits targeting either receptor-binding-domain (RBD) or spike (S) protein. The kits used in the present study include detection of IgM (WANTAI, Beijing Wantai Biological Pharmacy Ent.), IgA, IgG, and NAb (EUROIMMUN, Lubeck, Germany). Serologic testing was conducted at every time point, following the manufacturer’s protocols. The threshold for positivity of RBD-specific IgM is defined at an optical density of 0.105 OD at a wavelength of 450 nm (OD_450_), with a reference at 650 nm (OD_650_). For S1-specific IgA, semiquantitative results are derived by calculating the OD ratio using the samples and a provided standard. A positivity cut-off value of 1.1 OD ratio is established to interpret results. For S1-specific IgG, quantitative determination of binding antibody units (BAU/mL) is obtained using the standards provided by the manufacturer with a positivity cut-off value of 35.2 BAU/mL. Serum NAb activity against S1/RBD-Wuhan-Hu-1 SARS-CoV-2 (Wuhan wildtype) is measured in a surrogate Virus neutralization Test (sVNT) based on competitive ELISA. According to the manufacturer sheets, the sensitivity of these commercial ELISA kits ranges from 86.1% to 96.9%, and the specificity ranges from 98.3% to 99.8%.

### 4.2 Memory cells functionality through ELISpot

The functionality of memory B and T cells is analyzed using PBMCs stimulated against SARS-CoV-2 antigens in vitro. Commercial ELISpot assays (Mabtech, Sweden) quantify the production of RBD-specific IgG in memory B cells. The assays assess the production of cytokines, particularly interferon-gamma (IFN-γ) in memory T cells. Briefly, 2 x 10^5^ cells are incubated with SARS-CoV-2 peptides for 24 to 72 hours, and spot development is performed according to the manufacturer’s protocol. Unstimulated controls for each sample function as the internal reference, enabling a comparative analysis of stimulated changes using the individual’s baseline as a reference point. Spots are counted using an ImmunoSpot® analyzer and converted into spots forming unit per million cells (SFU/mL) for standardization. The collected data is analyzed to determine the frequency of cytokine-producing memory T cells and IgG-secreting memory B cells, as previously described [6].

### 4.3 B lymphocytes subsets response through flow-cytometric analyses

PBMCs were analyzed using flow cytometry to discern the frequency of distinct B cell subpopulations and their potential correlation with cell functionality. Flow cytometric analyses were designed in the present study to characterize the B cell response include cell viability (7-AAD), mature B cell lineage (CD19+ CD20+), transitional B cell (CD38 dim/- CD24+), antibody-secreting cell (CD38+ CD24- CD27+ CD20-), CD27 and IgD for atypical memory B cells, memory B cells, innate-like memory B cells, naïve B cells. Spike and RBD-specific SARS-CoV-2 markers, including BB515 Streptavidin BV421 Streptavidin for double spike+, BB515 Streptavidin BV421 Streptavidin for double spike+ with spike RBD+, and IgM with IgG to distinguish the primary and secondary response in the stimulated samples. Previous literature on cellular immunity response against SARS-CoV-2 were referred for the cells subset as previously describe [5,21]. A BD FASCLyric™ cell analyzer was be used to characterize the samples.

### 4.4 Cytokine profiling

In the subset of 45 selected participants, including the 5 unvaccinated control participants, cytokine levels from the supernatant of cells suspension are assessed using the Bio-Plex multiplex magnetic bead-based human cytokine assay (Bio-Rad, CA, USA) with the 27-Plex Screening Panel (#M500KCAF0Y). The Bio-Plex 200 Reader measures median fluorescence intensities, and each sample undergoes triplicate analysis. Data acquisition and analysis are performed using Bio-Plex Manager Software. Standard curves for each cytokine are established using the provided manufacturer’s standards according to manufacturer’s instruction and as described previously [22].

In this study, we focused on 27 unique cytokines which involves in inflammatory responses and initiate cellular immune signals: eotaxin, fibroblast growth factor (FGF), granulocyte colony-stimulating factor (G-CSF), interferon-γ (IFN-γ), IL-1β, interleukin 1 receptor antagonist (IL-1RA), IL-2, IL-4, IL-5, IL-6, IL-7, IL-8, IL-9, IL-10, IL-12(p70), IL-13, IL-15, IL-17A, interferon-γ-inducible protein (IP-10), monocyte chemoattractant protein-1 (MCP-1, MCAF), macrophage inflammatory protein-1α (MIP-1α), MIP-1β, platelet-derived growth factor-BB, regulated upon activation, normal T-cell expressed and presumably secreted (RANTES), tumor necrosis factor-α (TNF-α), and vascular endothelial growth factor-A (VEGF).

### 4.5 Proteome Profiling

Longitudinal serum samples were used to study the host proteomic response against COVID-19 vaccination and/or infection. During the protein preparation, high abundance proteins were depleted by a commercial kit, High Select™ Depletion Spin Columns (Thermo Scientific, USA), and protein digestion was performed using EasyPep™ MS Sample Prep Kits (Thermo Scientific, USA). Protein identification and quantification are performed using the untargeted Liquid Chromatography-Tandem Mass Spectrometry (LC-MS/MS) Q-TOF (Agilent Technologies 6520, USA) system. Samples were identified using data-independent acquisition mode. Spectra from each fraction are explored in the protein database, and the results are imported to generate a library to identify differentially expressed proteins (DEPs) that significantly vary between samples. The functional significance of DEPs is explored using bioinformatics tools, including Gene Ontology (GO) and KEGG pathway analysis following protocols of available study [23].

### 4.6 Data analysis and modeling plan

At the time of study conceptualization during the early phase of the pandemic, the initial objective was to compare the immune response between participants receiving only the BNT162b2 vaccine with no infection to those post-COVID-19 infection without vaccination, focusing on (1) measurement of serum-neutralizing antibodies, IgG, IgA, IgM response against SARS-CoV-2 for up to 12 months and (2) evaluation of the long term-memory B and T lymphocyte-mediated immune response. The inclusion of only BNT16b2 vaccine in the present study was informed by its predominant primary series vaccine used in Malaysia. Throughout the pandemic, the study analysis plans had to undergo continuous adjustments and adaptation to account for group overlap due to varying accumulation of SARS-CoV-2 exposures outside of the control of the study investigators, such as the implementation of the government’s vaccine rollout plan and the emergence of new more transmissible and immune-evasive variants that cause breakthrough infections. To accommodate these limitations, additional data on COVID-19 vaccination, booster doses, and COVID-19 infection histories were requested from the participants at the end of their last sampling timepoints. These data were collected from a local government-sponsored and mandated electronic record, the *MySejahtera* application available on all participants’ mobile devices to limit recall bias. The additional data were utilized to determine cross-exposure in participants, specifically examining the immune response against SARS-CoV-2 induced by either vaccination alone, infection alone, or a combination of both. Only participants who provided proof of vaccination certificate and diagnostic results from rapid test kits or PCR were included in the final analysis. Additionally, 4-fold increment increases in anti-SARS-CoV-2 S1 IgG over consecutive sampling timepoints (outside of the vaccination period) were monitored to account for asymptomatic infections. Participants in the VAC group with COVID-19 infection are grouped based on breakthrough infection (BTI) status. Plans to analyze these results are in progress and will be reported in future publications.

A complete-case analysis approach was chosen, wherein only participants who completed the entire follow-up period for blood sampling and provided fully completed Case Report Forms (CRFs) were included in the analysis. The assumption was that the excluded participants were represented randomly, indicating that the missing data was categorized as Missing Completely at Random (MCAR) data [24] and therefore the excluded data will not affect the statistical analyses.

Descriptive statistics were employed to summarize the characteristics of study participants and relevant variables, providing an overview of data distribution and cohort demographics. Age groups were categorized according to common life stages, which are group ≤ 30 years old (teenagers and young adults), 31-40 years old (adults), > 40 years old (late adults to seniors/elderly), respectively. Logistic regression will be used to assess the association between categorical outcomes (e.g., symptoms/AEFI severity, antibody waning status) and predictor variables (e.g., vaccination types, frequency of infection), while generalized estimating equation (GEE) models will be used to account for longitudinal data with repeated measurements, enabling the examination of changes in immune response markers and BTIs/reinfections over time. Correlation tests explore relationships between continuous variables, revealing the strength and direction of factor associations. Additionally, survival analysis is employed for time-to-breakthrough infection/reinfection, and cluster analysis identifies distinct participant subgroups based on immune response profiles or other characteristics such as the SARS-CoV-2 vaccination types and infection frequency. The analyses are performed using established statistical software (e.g., R, SPSS, GraphPad Prism).

### 5.0 FINDINGS TO DATE

Initially 184 SARS-CoV-2 naïve, SARS-CoV-2 anti-S1 IgG seronegative participants were enrolled into the VAC group in March 2021 and April 2021 before their scheduled BNT162b2 vaccination (Supplemental Figure 2). This was made possible by our university hospital’s designation as a vaccination distribution center, allowing us to enroll study participants even during MCOs. Starting in May 2021, however, there was a decrease in the number of SARS-CoV-2 naïve participants due to the Malaysian government’s mass vaccination campaign. This decline presented challenges in meeting our initial target of 200 participants for the VAC group.

In contrast, the enrollment for the COV group encountered a slowdown in the initial phase of the study, leading to an extension. This delay was prompted by unexpected restrictions imposed during the MCO implemented by the Malaysian government between March 2020 and December 2021 including limitations on gathering, mobility, and international travel, along with the closure of government, business, and educational institutions to prevent the spread of SARS-CoV-2. However, enrollment for the COV group experienced a significant increase in March 2022, coinciding with the emergence of the Omicron variant. Despite the challenges, we successfully enrolled 216 COV group participants with post-COVID-19 natural infection. An additional 16 participants were included to compensate for the participation numbers in the VAC group.

The decrease in retention rate started at Day-180 of sample collection (Supplemental Table 1). Through our observations and interactions with participants, the primary reason for study withdrawal was the resumption of their usual daily responsibilities during the transition from the pandemic to the endemic phase. Consequently, shifting priorities among participants led to a diminished interest in continuing their participation in the study. By the final follow-up on Day-360, the total retention rate throughout all sampling timepoints was 70% (n=280/400) which providing a sample at every single sampling timepoint.

### 5.1 Enrollment strategies

Given the ongoing COVID-19 pandemic during the study, we informed the participants of their serology test results to make them aware of their status against SARS-CoV-2. By sharing serology results, participants were likely motivated to remain engaged in the study, as they recognized the importance of continuous surveillance.

In addition, we also implemented a mobile follow-up service to enhance participant engagement especially during the MCOs. The research team contacted participants to schedule home visits for sample collection to ensure a higher retention rate. Overall, from our observation, keeping participants informed about the study’s progress, providing timely updates, and addressing their concerns were the key factors in building strong trusting relationships and rapport with the participants. These strategies strengthened our enrollment and retention efforts, fostering an engaged cohort. This approach has not only empowered individuals from various backgrounds to participate actively but also contributed to the overall success and validity of our research study.

### 5.2 Modification of the study designs

A limitation within our study design arises from the early conceptualization during the initial stages of the pandemic, resulting in cross-exposure between the initial study groups. Consequently, the distinction between breakthrough infections in the vaccination-only (VAC) group and vaccinations in the infection-only (COV) group was not effectively addressed. This limitation hindered the ability to exclusively isolate the immune response from either VAC or COV throughout the entire blood sampling period. Furthermore, there is a possibility of underreported data on previous infection history due to the potential occurrence of asymptomatic infections. Another important limitation is that we only gathered data on symptoms and adverse events following immunization (AEFI) during the enrollment phase, without continuing to track during the breakthrough infections and reinfections in the later stages of the study.

We acknowledged the potential confounding effects from individual behaviors and personality traits when assessing the influence of vaccination timing on SARS-CoV-2 breakthrough infection risk. Variability in risk perception and health-related behaviors between early vaccine recipients and those delaying vaccination, alongside socioeconomic disparities and differential access to healthcare, present significant challenges in analysis. Therefore, we will perform multivariate analyses and sensitivity analyses to assess the robustness of findings across different exposures, while using regression modeling control for confounding variables.

### 5.3 Cross-exposure of SARS-CoV-2 vaccination and infection from initial groupings

With the emergence of breakthrough infections and reinfections, we reconsidered the initial stratification of VAC and COV groups due to cross-exposure between the 2 groups. Supplemental Figure 3 shows an overlapped graph depicting the sampling period of LONGTONG-SARS2 study and the national SARS-CoV-2 variants genomic surveillance data in Malaysia. We expected significant hybrid immunity, arising from both natural infections followed by vaccination and vice-versa, to be present in both participant groups amidst the emergence of highly transmissible variants and the high number of COVID-19 cases in combination with mass country-wide vaccination campaigns (Supplemental Figure 3, Supplemental Figure 4, Supplemental Table 2).

During the peak of Delta and Omicron circulation, when these variants constituted 75% of the COVID-19 cases in Malaysia (Hodcroft, 2023), we enrolled 67 and 222 participants, respectively. Those participants were likely infected with those variants based on the dominant circulating variants during the enrollment (Supplemental Figure 4). However, the present study did not involve sequencing respiratory samples. Instead, we inferred the participants’ likely infection variant based on contemporary national genomic sequencing data [25].

During the peak of the Delta variant, 16.8% of all participants (n=67/400) were identified, with 23.6% (n=51/216) from the COV group and 8.7% (n=16/184) from the VAC group. Similarly, during the peak of the Omicron variant, 55.5% of all participants (n=222/400), with 67.1% (n=145/216) in the COV group and 41.8% (n=77/184) in the VAC group.

Cross-exposure of SARS-CoV-2 vaccination and infection was observed from the initially stratified participants throughout the 12-month sampling period in the present study (Supplemental Table 2). Almost two-thirds (68.8%) of all study participants (n=275/400) encountered breakthrough infections. Notably, within the VAC group, more than half (53.3%) experienced breakthrough infections (n=98/184). Furthermore, almost every participant (98.6%) in the COV group (n=213/216) had received a full primary series COVID-19 vaccination due to mass vaccination campaigns implemented by the Malaysian government to ensure a high vaccination rate. Detailed demographic data of this subset population can be referred to the Table 1 and Table 2.

**Table 1.** Distribution of sociodemographic factors by sex and age groups according to initial participant grouping, VAC versus COV. Abbreviations: VAC = SARS-CoV-2-naïve prior to scheduled BNT162b2 primary series vaccination; COV = recently recovered from COVID-19 infection.

|  | VAC | COV | Total | X<sup>2</sup> Stat (df) | p-value |
| --- | --- | --- | --- | --- | --- |
| Sex |  |  |  |  |  |
| Male | 60 (42.5%) | 45 (35.7%) | 105 (39.3%) | 1.30 (1) | 0.253 |
| Female | 81 (57.4%) | 81 (64.3%) | 162 (60.7%) |  |  |
| Age group |  |  |  |  |  |
| ≤ 30 | 17 (12.1%) | 37 (29.4%) | 54 (20.2%) | 15.11 (2) | 0.001 |
| 31-40 | 68 (48.2%) | 59 (46.8%) | 127 (47.6%) |  |  |
| > 40 | 56 (39.7%) | 30 (23.8%) | 86 (32.2%) |  |  |
| Total | 141 (52.8%) | 126 (47.2%) | 267 (100%) |  |  |

**Table 2.** Distribution of sociodemographic factors of sex and age group by revised study group.

|  | E/VAC(BNT)/COVID-ve | E/VAC(BNT)/COVID+ve | VAC(BNT)/COVID+ve/E | VAC(any)/COVID+ve/E | COVID+ve/E/VAC(any) | Total | X<sup>2</sup> Stat (df) | p-value |
| --- | --- | --- | --- | --- | --- | --- | --- | --- |
| **Sex** |  |  |  |  |  |  |  |  |
| Male | 27 (26%) | 30 (29%) | 22 (21%) | 14 (13%) | 12 (11%) | 105 (39%) | 4.66 (4) | 0.324 |
| Female | 25 (15%) | 55 (34%) | 40 (25%) | 25 (15%) | 17 (11%) | 162 (61%) |  |  |
| **Age group** |  |  |  |  |  |  |  |  |
| <=30 | 7 (13%) | 10 (19%) | 12 (22%) | 18 (33%) | 7 (13%) | 54 (20%) | 26.94 (8) | 0.001 |
| 31-40 | 24 (19%) | 42 (33%) | 35 (28%) | 15 (12%) | 11 (9%) | 127 (48%) |  |  |
| >40 | 21 (24%) | 33 (38%) | 15 (17%) | 6 (7%) | 11 (13%) | 86 (32%) |  |  |
| **Total** | **52 (20%)** | **85 (32%)** | **62 (23%)** | **39 (15%)** | **29 (11%)** | **267 (100%)** |  |  |

### 5.4 Participants’ demographics and study groups restructuring

Of the 400 participants, 70% (n=280) completed the blood sampling follow-ups (Supplemental Table 1). Among them, 66.8% (n=267) completed the follow-ups and CRF. These 267 were included for the final analyses.

Of the 267 participants, 52.8% (n=141) are from the VAC group, while the remaining 47.2% (n=126) belonged to the COV group (Table 1). Demographic characteristics revealed a balanced sex distribution, with 39.3% male and 60.7% female participants (p=0.253). Age distribution showcased a significant difference across age groups (p=0.001), with 20.2%, 47.6%, and 32.2% of participants falling into age groups ≤ 30 years old, 31-40 years old, and > 41 years old, respectively. Median age of participants was 36 years old (x̅=37, IQR=9 to 73).

In the revised study participant grouping, our study delineated five distinct subgroups. These included “E/VAC(BNT)/COVID-ve”, “E/VAC(BNT)/COVID+ve”, “VAC(BNT)/COVID+ve/E”, “VAC(any)/COVID+ve/E”, and “COVID+ve/E/VAC(any)” (Figure 3). Where the acronym is defined as “*E*” is Enrolled; “*VAC(***)*” is vaccination type either BNT162b2 or any vaccinations; “*COVID*ve*” is the status of COVID-19 infection either positive or negative. Forward slashes in the revised group names separate distinct events in chronological order.

**Figure 3.**
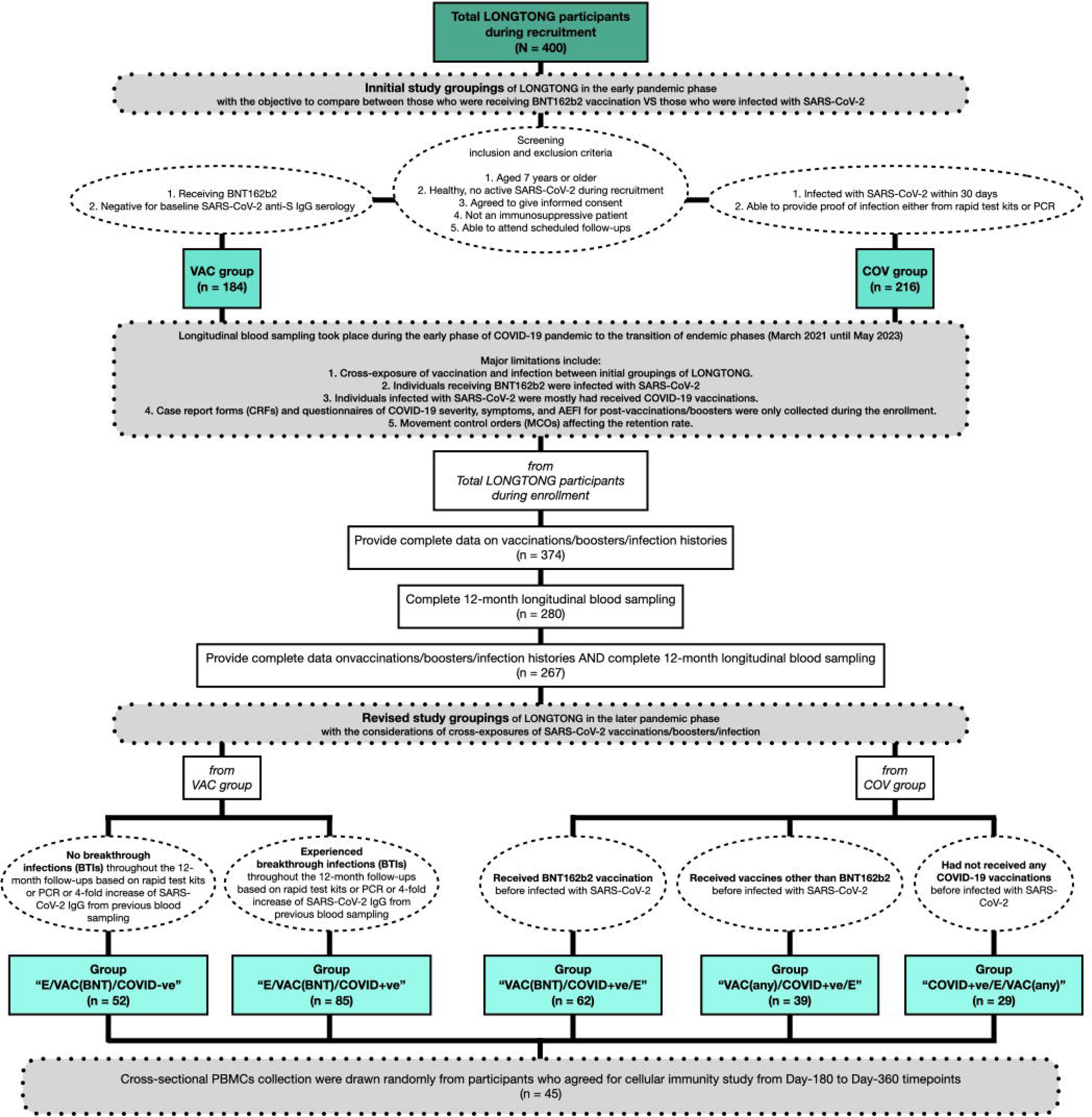
Flow chart of group stratification changes from initial study groupings to revised study groupings to accommodate the cross-exposures in the participants immune response.

In detail, the five distinct groups within our study cohort, each defined by their vaccination history and COVID-19 infection status:

1. "E/VAC(BNT)/COVID-ve" = this group comprised participants who were enrolled, received the primary series of BNT162b2 vaccination, and did not experience a breakthrough COVID-19 infection.
2. "E/VAC(BNT)/COVID+ve" = this group comprised participants who were enrolled, received the primary series of BNT162b2 vaccination, and had documented breakthrough COVID-19 infection.
3. "VAC(BNT)/COVID+ve/E" = this group comprised participants who received the primary series of BNT162b2 vaccination prior to enrollment in the study, later had documented breakthrough COVID-19 infection, and subsequently enrolled the study.
4. "VAC(any)/COVID+ve/E" = this group comprised participants vaccinated with a primary series of COVID-19 vaccination (other than BNT162b2), later had documented breakthrough COVID-19 infection, and subsequently enrolled in the study.
5. "COVID+ve/E/VAC(any)" = this group comprised participants who had documented COVID-19 infection prior to study enrollment and subsequently received a primary series of COVID-19 vaccination (other than BNT162b2).

The results of the Chi-square test indicate that the distribution of sex and age groups in the revised study group is comparable to that in the initial study groups (Table 2). Median age of participants in the revised group was 36 years old (x̅=38, IQR=15 to 65). There were no significant differences in sex (p-value=0.324); however, a significant difference was observed among age groups (p-value=0.001).

Additionally, for the cross-sectional PBMCs collection, we will conduct a comparative analysis of cellular immune responses among individuals in both homologous and heterologous vaccination groups, further stratifying them based on breakthrough infection occurrence (Supplemental Table 3). The cellular responses within these vaccinated groups will be compared with those of an unvaccinated control group.

### 6.0 FUTURE DIRECTIONS

Studies are planned or ongoing examining the kinetics of antibody production post-SARS-CoV-2 vaccination and infection. Preliminary analysis indicates variations among the revised study groups, prompting further investigation into the factors influencing the rate of decline and persistence of antibody levels over time in response to breakthrough infection and/or reinfection. Furthermore, in our exploration of the potential correlates of protection, we will examine the specific antibody, memory B cell, and proteomic response that best correlate with durability and functionality of immune response against SARS-CoV-2. These analyses are pending. Additional investigations will examine how previous exposures to SARS-CoV-2, whether through vaccination or infection, impact adaptive immune responses during subsequent exposures to elucidate the role of memory B cells and memory T cells in sustaining long-term immunity. Taken together, the cohort from the present study will be leveraged to delve into a focused exploration of the heterogeneity of SARS-CoV-2 immune responses arising from diverse vaccination regimens and antigen exposures.

### 7.0 ETHICS, COLLABORATION, AND DISSEMINATION

This study is strictly adhered to the ethical principles outlined in the World Medical Association Declaration of Helsinki. Ethical approval was obtained from Universiti Malaya Medical Centre (MREC-UMMC SID: 2021226-9886), ensuring compliance with all relevant federal regulations governing the protection of human subjects. All study personnel hold certifications in Good Clinical Practice. The study is supported by the U.S. Naval Medical Research Unit INDO PACIFIC (NAMRU INDO PACIFIC) and will adhere to the NAMRU INDO PACIFIC regulations for publication in open-access journals and data management.

The research team welcomes opportunities for potential research collaborations. The data from this study can be obtained upon request from the corresponding author, SAB. Permission to access data and analytical files is contingent upon approval from the relevant research ethics committees and the data custodian. The LONGTONG-SARS2 biological materials and data custodians are the Tropical Infectious Diseases Research and Education Centre (TIDREC) and the U.S. Naval Medical Research Unit INDO PACIFIC (NAMRU INDO PACIFIC).

Key study findings will be disseminated through various channels to ensure widespread and transparent communication. Results will be published in journals for the scientific community. Furthermore, the findings will be presented at national and international conferences, facilitating knowledge exchange and fostering collaboration with researchers worldwide. Notably, the study team is committed to communicating the results to the study population. By ensuring direct and accessible communication, participants will be informed on the progress and outcomes of the present study in which they have actively participated.

## Data Availability

All data produced in the present study are available upon reasonable request to the authors

## Acknowledgment

We acknowledge the dedication and commitment of the voluntary study participants without whom this project would not have been possible.

## Authors’ contribution

SAB had full access to all the study data and takes responsibility for the integrity of the data and the accuracy of the data analysis. Study concept and design: SAB, JJ, RH, NCK. Participant enrollment and data collection: NCK, JJ. Sample processing: NCK. Laboratory work: NCK. Data analysis and interpretation: NCK. Original drafting of manuscript: NCK. Editing and revision of manuscript: NCK, JJ, HY, HN, AL, RH, SAB. Study coordination: SAB, JJ, HY, HN, AL, RH, NCK. All authors read and approved the final article.

### Disclaimer

The views expressed in this article are those of the authors and should not be construed to reflect the official policy or represent the positions of the U.S. Department of the Navy, Department of Defense, nor the United States Government. LT Huy C. Nguyen, MC, USN NAMRU-IP; LCDR Robert D. Hontz, MSC, NAMRU-IP; CAPT Andrew G. Letizia, MC, USN NAMRU-IP are military service members. This work was prepared as part of their official duties. Title 17 U.S.C. 105 provides that ‘copyright protection under this title is not available for any work of the United States Government.’ Title 17 U.S.C. 101 defines a U.S. Government work as work prepared by a military service member or employee of the U.S. Government as part of that person’s official duties.

