## Supplemental for "Cohort Profile: Immune Responses to SARS-COV-2 Vaccination and Infection in a Longitudinal Sampling Amidst the COVID-19 Pandemic (LONGTONG-SARS2) in Malaysia"

**SUPPLEMENTAL TABLES AND FIGURES**

**Supplemental Table 1.** Retention rate of participation in the long-term follow-ups in LONGTONG-SARS2 study.

| **Group** | **Timepoint** | **Retention rate** |
| --- | --- | --- |
| VAC  (n=184) | 1. Pre-vaccination  2. Day-7 post first dose  3. Day-14 post first dose  4. Day-7 post second dose  5. Day-14 post second dose  6. Day-90 post first dose  7. Day-180 post first dose  8. Day-270 post first dose 9. Day-360 post first dose | 100% (n=184/184)  100% (n=184/184)  100% (n=184/184)  100% (n=184/184)  100% (n=184/184)  100% (n=184/184)  79.9% (n=147/184)  78.8% (n=145/184)  78.8% (n=145/184) |
| COV  (n=216) | 1. Day-14 post syndrome onset  2. Day-30 post syndrome onset  3. Day-60 post syndrome onset  4. Day-90 post syndrome onset  5. Day-180 post syndrome onset  6. Day-270 post syndrome onset  7. Day-360 post syndrome onset | 100% (n=216/216)  100% (n=216/216)  100% (n=216/216)  100% (n=216/216)  80.6% (n=174/216)  76.4% (n=165/216)  62.5% (n=135/216) |

**Supplemental Table 2.** Cross-exposure of LONGTONG-SARS2 study participation in the initial study groupings (VAC and COV) either through breakthrough infection (BTI) and/or vaccination among both groups.

| **Study group** | | **Received COVID-19 vaccination** | | **Breakthrough infections** |
| --- | --- | --- | --- | --- |
| VAC | 100.0% (n=184/184) | | 53.3% (n=98/184) | |
| COV | 98.6% (n=213/216) | | 81.9% (n=177/216) | |
| Total | 99.3% (n=397/400) | | 68.8% (n=275/400) | |

**Supplemental Table 3.** Distribution of Peripheral Blood Mononuclear Cells (PBMCs) samples selected for analysis according to COVID-19 vaccine schedules and frequency of COVID-19 infection. BNT/BNT denotes BNT162b2 COVID-19 vaccine (Comirnaty, Pfizer-BioNTech) for both primary and booster doses. ChAd/ChAd denotes ChAdOx1 nCoV-19 (ChAd) COVID-19 vaccine (Vaxzevria, AstraZeneca) for both primary and booster doses. CoVa/CoVa denotes CoronaVac COVID-19 vaccine (Sinovac) for both primary and booster doses. ChAd/BNT denotes a ChAd vaccine for the primary dose and a BNT vaccine for the booster dose. CoVa/BNT denotes a CoronaVac vaccine for the primary dose and a BNT vaccine for the booster dose.

| **COVID-19 vaccine schedule** | **Frequency of COVID-19 infection** | | | ***Total*** |
| --- | --- | --- | --- | --- |
|  | **0** | **1** | **2** |  |
| **Homologous (n=30)**  **(Booster type identical to primary series)** |  |  |  |  |
| BNT/BNT | 3 | 5 | 2 | ***10*** |
| ChAd/ChAd | 2 | 7 | 1 | ***10*** |
| CoVa/CoVa | 2 | 6 | 2 | ***10*** |
| **Heterologous (n=15)**  **(Booster type different from primary series)** |  |  |  |  |
| ChAd/BNT | 0 | 5 | 1 | ***6*** |
| CoVa/BNT | 2 | 6 | 1 | ***9*** |
| **No COVID-19 vaccination [Unvaccinated control] (n=5)** | 1 | 4 | 0 | ***5*** |
| ***Total*** | ***10*** | ***33*** | ***7*** | ***50*** |

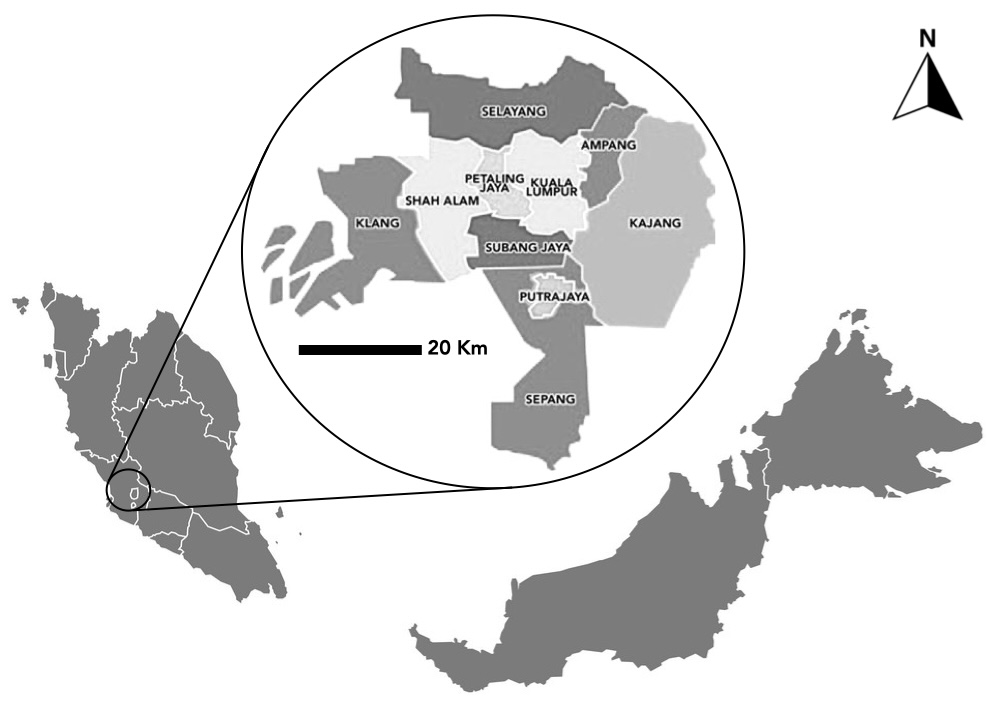

**Supplemental Figure 1.** Malaysia map with the greater Kuala Lumpur location (figure inset).

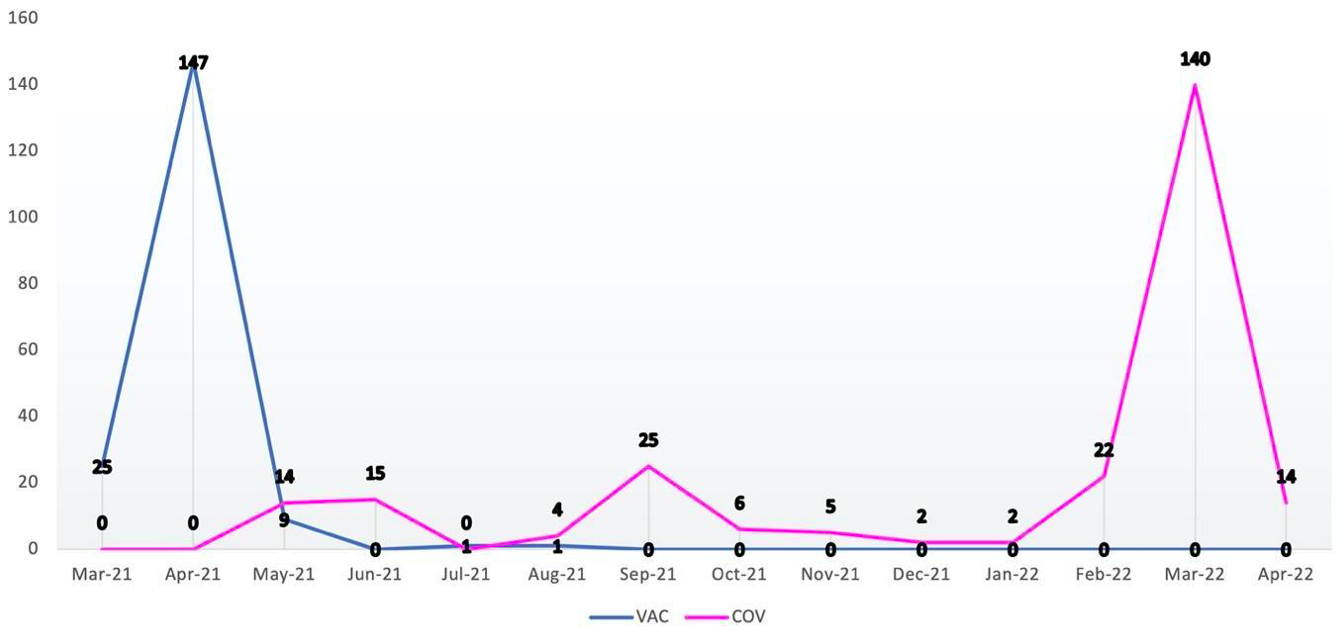

**Supplemental Figure 2.** Distribution pattern of LONGTONG-SARS2 study enrolment.

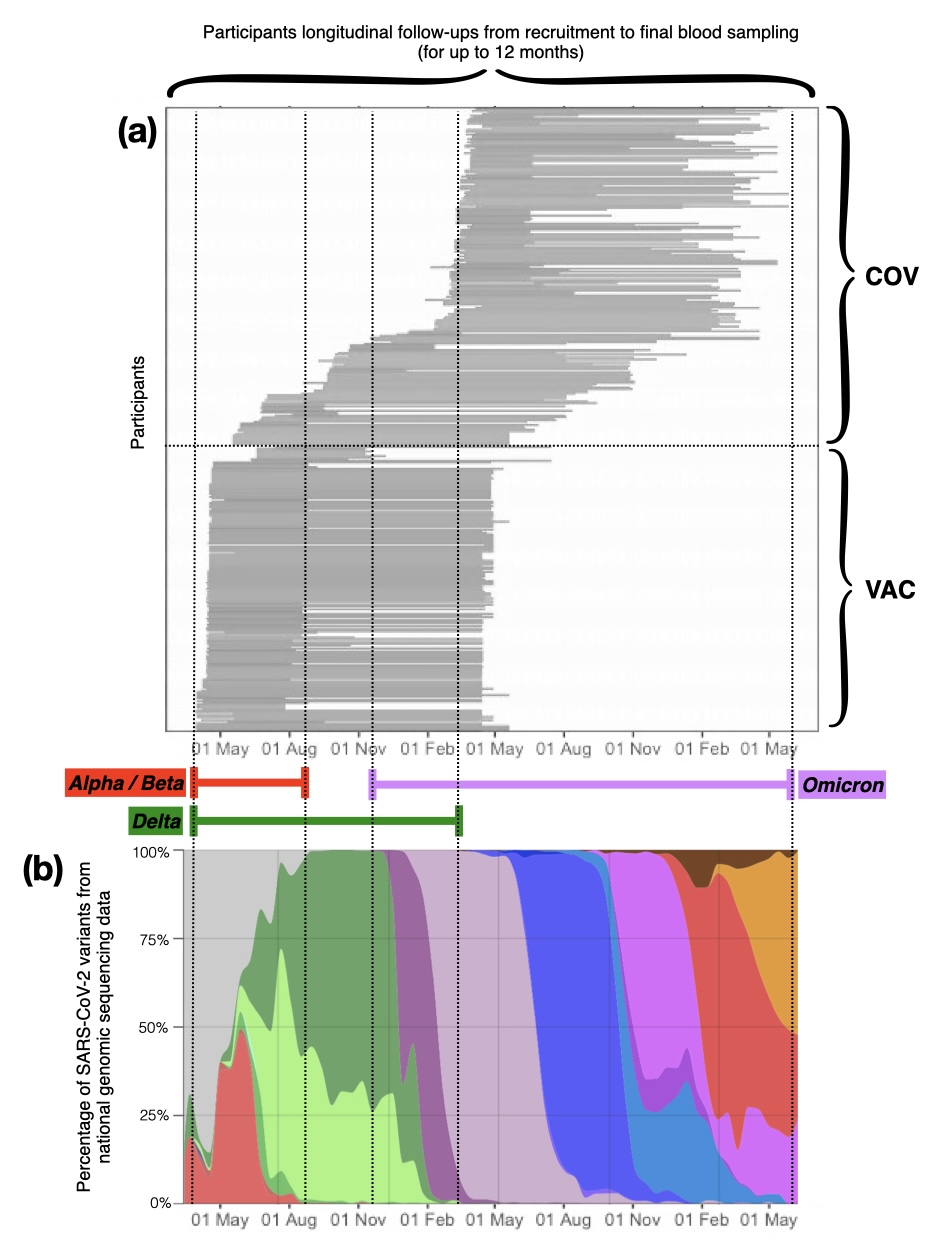

**Supplemental Figure 3.** Longitudinal blood sampling from March 2021 to May 2023 with the SARS-CoV-2 variants during the sample collection. The dotted lines show the overlap of blood sampling period to the predominant SARS-CoV-2 variants from the national genomic sequencing data. (a) Sampling timeline for up to 12 months. Each horizontal line represents longitudinal follow-ups for one participant. (b) Percentage of SARS-CoV-2 variants distribution in Malaysia during the sampling period.

**Supplementary Figure 3(b) was sourced from Hodcroft (2023): CoVariants (Link: https://covariants.org/per-country?country=Malaysia)*

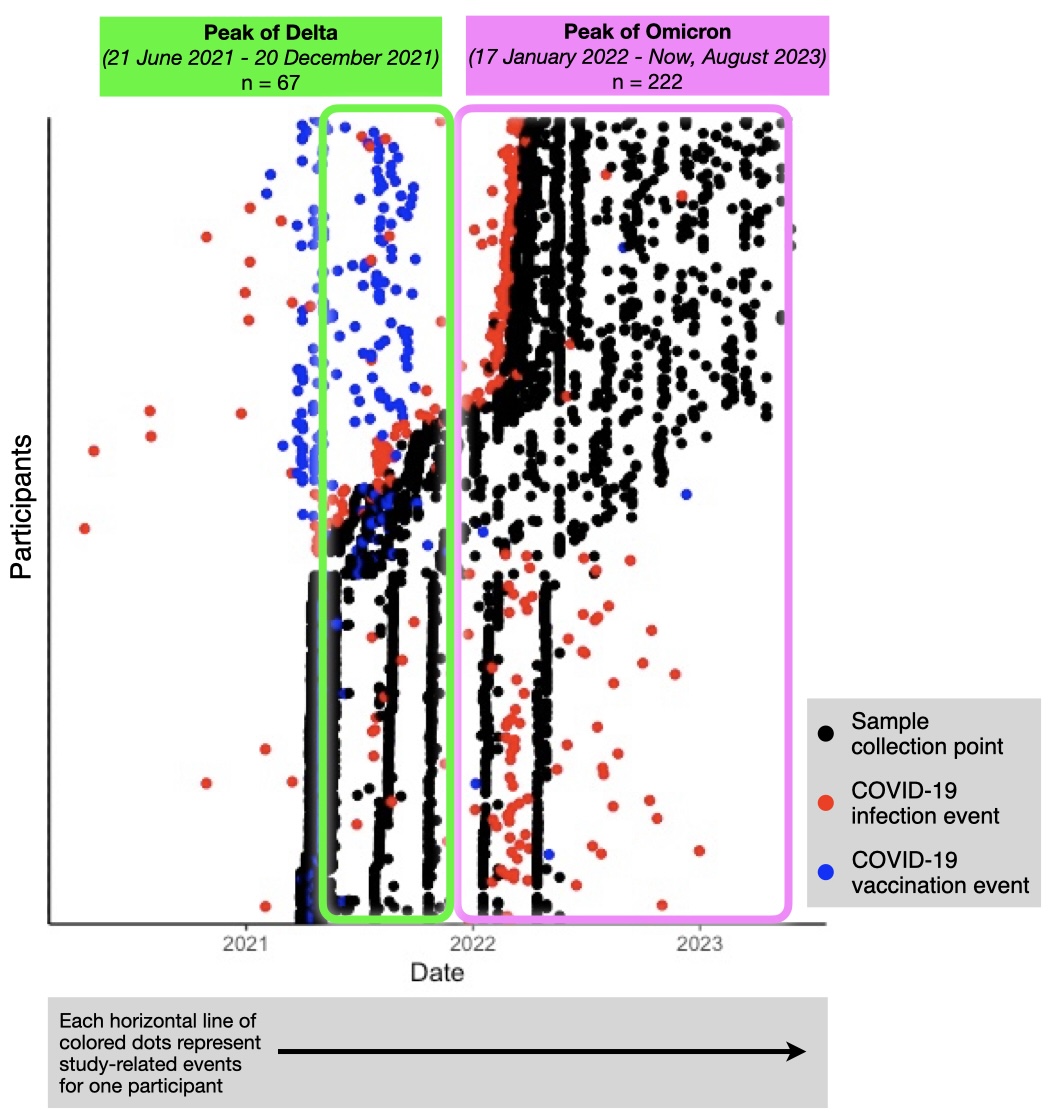

**Supplemental Figure 4.** Participants from total samples (N = 400) were exposed to SARS-CoV-2 during Malaysia's peak of Delta and Omicron variants. The sample collection points, COVID-19 vaccination and infection events from participants (if available) are shown for each participant.
